# Predicting hospital readmissions in patients receiving novel-dose Sacubitril/Valsartan therapy: A competing-risk, causal mediation analysis

**DOI:** 10.1101/2023.02.08.23285680

**Authors:** Changchun Hou, Xinxin Hao, Ning Sun, Xiaolin Luo, Luyu Wang, Enpu Yang, Zhichun Gao, Ling Chen, Zebi Wang, Yun Cui, Jing Zhong, Juhao Yang, Xi Liu, Zhexue Qin

## Abstract

**Background:** The effects of novel-dose Sacubitril/Valsartan (S/V) in patients with heart failure (HF) in the real world have not been adequately studied. We examined the risk for all-cause re-admission in the patients with HF taking novel-dose S/V and the possible mediator role of left ventricular reverse remodeling (LVRR).

**Methods and Results:** There were 464 patients recruited from December 2017 to September 2021 in our hospital with a median follow-up of 660 days (range, 17-1494). Model 1 and 2 were developed based on the results of univariable competing risk analysis, least absolute shrinkage and selection operator approach, backward stepwise regression and multivariable competing risk analysis. The internal verification (data-splitting method) indicated that Model 1 had better discrimination, calibration, and clinical utility. The corresponding nomogram showed that patients aged 75 years and above, or taking the lowest-dose S/V (≤50mg twice a day), or diagnosed with ventricular tachycardia, or valvular heart disease, or chronic obstructive pulmonary disease, or diabetes mellitus were at the highest risk of all-cause readmission. In the causal mediation analysis, LVRR was considered as a critical mediator that negatively affected the difference of novel-dose S/V in readmission.

**Conclusions:** A significant association was detected between novel-dose S/V and all-cause readmission in HF patients, in part negatively mediated by LVRR. The web-based nomogram could provide individual prediction of all-cause readmission in HF patients receiving novel-dose S/V. The effects of different novel-dose S/V are still needed to be explored further in the future.

## Introduction

Heart failure (HF) was a complex clinical syndrome that negatively impacted quality of life, and placed a huge and costly burden on global healthcare system probably causing by high readmission rates^1-3^. As the first agent of angiotensin receptor-neprilysin inhibitor (ARNI), Sacubitril/Valsartan (S/V) has been proven to significantly reduce all-cause readmission and mortality of patients diagnosed with HF with reduced ejection fraction (HFrEF)^4-7^. In clinical practice, however, many patients cannot achieve standard dose (97/103 mg twice a day [b.i.d.]), or even the lowest approved dose (24/26 mg b.i.d.) due to several factors (i.e., hypotension, hyperkalemia, renal dysfunction)^8-10^. Given novel-dose S/V (below the standard dose) would be used to treat HF patients in the real world, the clinical effects were of particular concern to clinicians.

A clear and early benefit of S/V was to reduce hospital readmissions of HF patients due to any causes^11^. McMurray et al. and Desai et al. pointed out that S/V was superior to enalapril in improving HF patient outcomes^4,5^. Lately, Carnicelli et al. reported that patients with higher adherence to S/V showed a significant reduction in all-cause readmission at 3 months or 1 year^12^. Unfortunately, there remained poor understanding of novel-dose S/V^13^. It would be of importance and interest to estimate the effects of novel-dose S/V on patients’ readmissions.

Moreover, left ventricular reverse remodeling (LVRR) has the potential to play a role in the beneficial effects of S/V^14-16^. Specially, some previous studies have demonstrated a strong favorable effect conferred by S/V on LVRR^15,17,18^, meanwhile some have reported a positive association between LVRR and clinical outcomes of HF patients^19,20^. It was well known that LVRR was pivotal to the progression of HFrEF patients^21-23^, while far fewer studies have examined its mediation effects on the relationship between S/V and patients’ outcomes.

Herein, this study would estimate the effects of novel-dose S/V on hospital readmissions, construct a risk prediction model, as well as investigate the possible role of LVRR. The findings of our study would hopefully provide crucial insights into the novel-dose S/V, and assist clinicians in selecting treatment options for patients to improve therapeutic outcomes.

## Methods

### Data sources

The present study complied with the Declaration of Helsinki and was approved by the ethics committee of Xinqiao Hospital, Army Medical University (Third Military Medical University) in Chongqing, China. We retrieved the data of 2424 patients diagnosed with HF from Xinqiao Hospital from December 29st, 2017 to September 17st, 2021. Patients would be included in our study if the following criteria were met: (1) age ≥18 years, (2) LVEF <50%, (3) an echocardiography was performed as historical data, and another would be performed as a comparison, (4) receiving novel-dose S/V, and would not discontinue therapy during the study period. Exclusion criteria included:(1) underwent CRT or cardiac transplantation therapy before or during the follow-up, (2) loss of follow-up. The flowchart of patients’ selection was shown in Figure 1.

**Figure 1.**
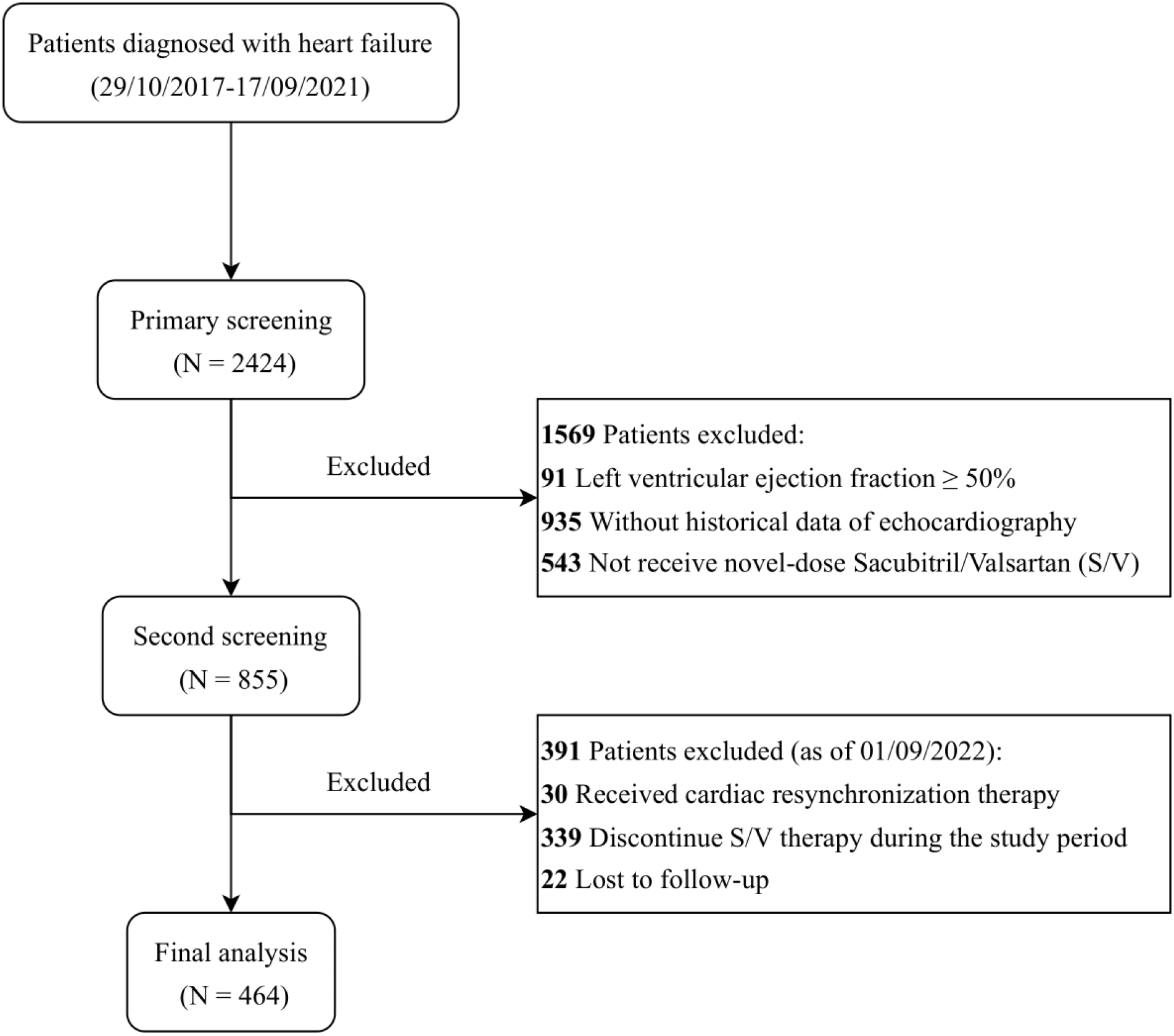
The flowchart of patients’ selection

### Data collection

HF-associated raw data was to be mainly collected from the medical records as following: (1) demographic characteristics, i.e., identification number, sex, date of birth, height, weight, etc., (2) comorbidities, i.e., dilated cardiomyopathy (DCM), myocardial infarction (MI), hypertension, coronary heart disease (CHD), valvular heart disease (VHD), chronic obstructive pulmonary disease (COPD), diabetes mellitus (DM), severe renal impairment (SRI), (3) electrocardiogram, i.e., premature atrial contractions (PACs), premature ventricular contractions (PVCs), ventricular tachycardia (VT), (4) echocardiography, i.e., left ventricular ejection fraction (LVEF), left ventricular end-diastolic diameter (LVEDD).

All included patients were followed up about 1 year (as of September 1st, 2022). The primary outcome was all-cause readmission, and other outcomes of interest was all-cause death.

### Statistical analysis

Given the study aimed to estimate the effects of different novel-dose levels, all patients were divided into three groups based on the time-weighted average dose^24^, that was, lowest dose (≤50mg b.i.d.), lower dose (50-<100mg b.i.d.) and low dose (100+mg b.i.d.), respectively. Then we split the dataset into training dataset and testing dataset with the ratio of 6:4 stratified by dose groups, which were used to develop predictive model (nomogram) and assess the performance, respectively. All statistical analyses were performed with R-4.2.1.

In order to correctly estimate the probability of the primary outcome of interest (i.e., all-cause-readmission), an extension of survival analysis, competing risk analysis, would be employed to predict all-cause readmission associated with different novel-dose S/V, which took into account the fact that competing outcomes (i.e., all-cause death) could prevent the occurrence of primary endpoint^25^. Firstly, cumulative incidence function (CIF) was used for showing the probability of clinical outcomes (all-cause readmission or death) in those receiving novel-dose S/V over follow-up time, and Gray’s Test was employed to test the equality of CIF curves in novel-dose subgroups. Secondly, univariable competing risk analysis was applied to explore the associations between covariates (i.e., demographic characteristics, comorbidities, electrocardiogram) and all-cause readmission, and those with a P-value < 0.2 would be identified as the potential prognostic variables^26^. If one of dummy variables was significant or met the inclusion criteria, the variable should be remained and considered as a unit in a model^27^. Thirdly, in order to avoid missing covariates that were clinically significant, two variable selection methods would be combined to determine the potential predictors for multivariable competing risk analysis, that was, least absolute shrinkage and selection operator (LASSO) approach with 5-fold cross validation and multivariable regression using stepwise backward selection based on Akaike’s Information Criterion (AIC)^1^. Fourthly, our study would produce two potential models with different predictors using multivariable competing risk analysis, that was, Model 2 (statistically significant variables with P-value < 0.05, and marginally significant with 0.05 < P-value < 0.1 according to the results of LASSO and AIC), Model 2 (statistically significant variables with P-value < 0.05), respectively. Meanwhile, two nomograms based on the multivariable regression models would be constructed to predict all-cause readmission of individual with HR receiving novel-dose S/V by weighting prognostic factors. Fifthly, a precise prognostic nomogram would be selected through evaluating the performance, including discrimination (i.e., concordance index (C-index) based on bootstrap, time-dependent area under the curve (AUC)), calibration (i.e., calibration curve), and net benefit (i.e., decision curve analysis (DCA)). C-index and time-dependent AUC values exceed 0.7 suggested a reasonable estimation^28^. Accordingly, a web-based dynamic nomogram would be built as a decision-making tool.

Moreover, non-linear causal mediation analysis was used to further clarify the association between novel-dose S/V and all-cause readmission through LVRR as the potential mediation (Figure S1). Specially, to promote LVRR was defined as an improvement ≥10% in left ventricular ejection fraction (LVEF) accompanied by a reduction of left ventricular end-diastolic diameter (LVEDD) ≥ 10%, or a LVEF increase ≥10% with at least 6-month follow-up^29-31^. First and foremost, S/V dosage was considered as a continuous variable to assess the significance of omnibus mediation effect. Then categorical novel-dose S/V was used to estimate the relative mediation effect, of which the lowest was the reference. Note that the estimated effect values were measured in terms of the log of hazards (odds of having all-cause readmission), which could be interpreted as the average differences in the log(hazards) between different levels of novel-dose S/V. Mediation analysis commonly decomposed the total effect into direct effect and indirect effect through mediators^32-33^, as shown in Figure S1.

## Results

### Study population

A total of 464 patients with a median follow-up of 660 days (interquartile range (IQR):17-1494) were entered into our dataset, which comprised 122, 173 and 169 receiving lowest-, lower- and low-dose S/V, respectively. 278 patients (median follow-up 646 days, IQR: 21-1487) and 186 patients (median follow-up 687 days, IQR: 17-1494) were divided into the training dataset (model development) and testing dataset (model performance), respectively. The baseline characteristics of these patients were summarized in Table S1. In general, a greater proportion of these three groups of patients (lowest-, lower- and low-dose S/V) were male, or those aged <55 years old, or diagnosed with DCM or VHD.

### Novel-dose related cumulative incidence

Gray’s Test showed that there was statistically significant difference in all-cause readmission (P-value < 0.001) across the three novel-dose groups, whereas no significant difference in all-cause death (P-value = 0.307). As illustrated in Figure 2, the estimated cumulative incidences of all-cause readmission of patients receiving the lowest-dose S/V were 4%, 10%, 15%, 25%, 30%, 43% at 30 days, 3 months, 6 months, 1 year, 2 years, 3 years, respectively. The corresponding estimates for patients taking lower-dose S/V were 0%, 1%, 2%, 4%, 11%, 18%, and for those with low-dose S/V were 0%, 2%, 3%, 4%, 6%, 8%, respectively.

**Figure 2.**
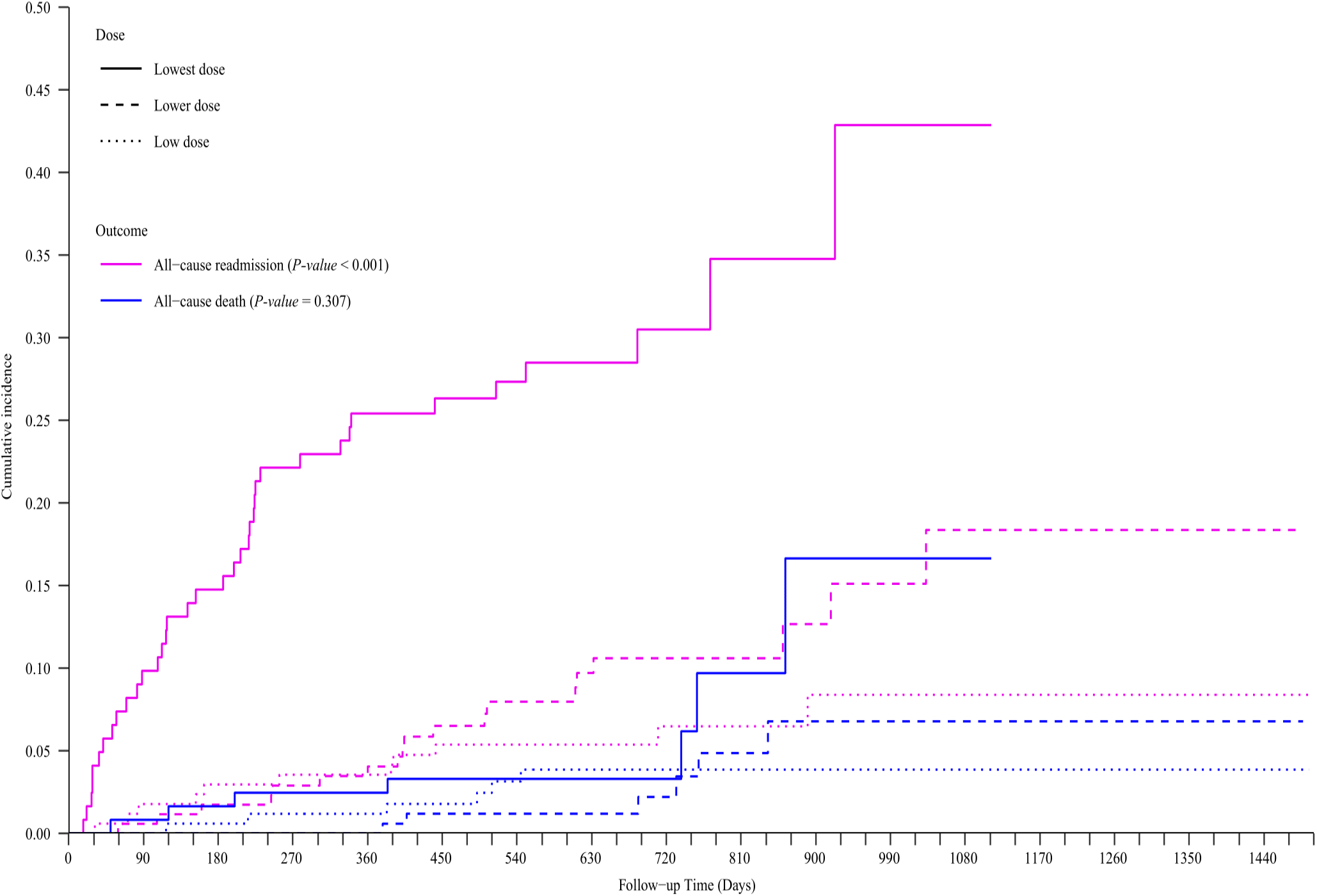
Cumulative incidence curves for all-cause readmission and death stratified by novel-dose Sacubitril/Valsartan on the basis of all dataset.

**Figure 3.**
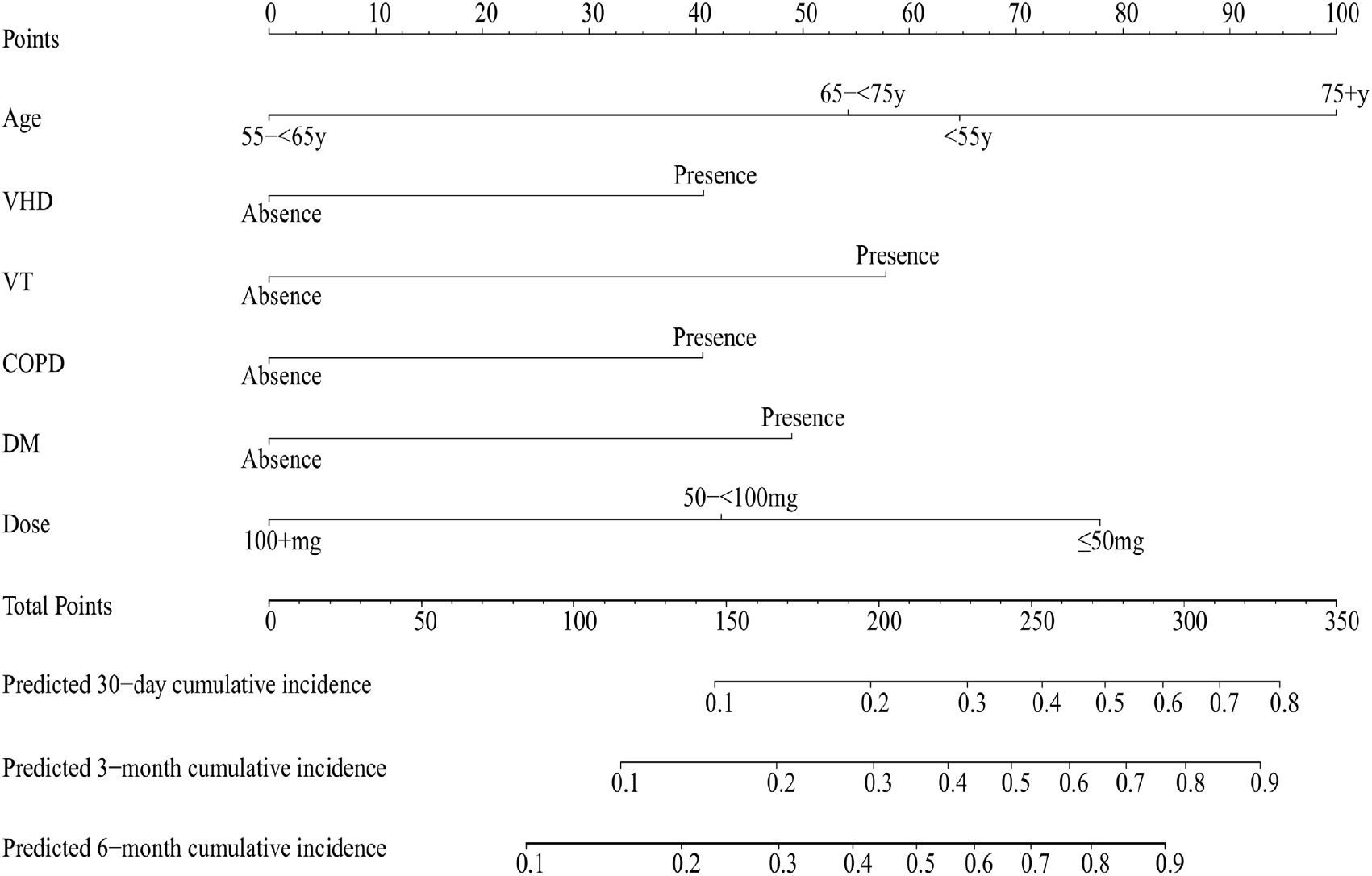
Nomogram for predicting 30-day, 3-month and 6-month cumulative incidence of patients with heart failure (in accordance with Model 1 developed by training dataset). Abbreviations: VHD, valvular heart disease; VT, ventricular tachycardia; COPD, chronic obstructive pulmonary disease; DM, diabetes mell

### Risk prediction model

Univariate competing risk analysis (Table 1) revealed that age, BMI, VHD, PACs, PVCs, VT, COPD, DM, SRI, dose were identified as the potential prognostic factors on all-cause readmission of HF patients. Then LASSO approach (Figure S2) and stepwise backward elimination method (Table S2) suggested that age, VT, DM and dose were significantly associated with patients’ readmission, while VHD and COPD were marginally significant variables. As shown in Table 1, our study constructed two models to predict primary outcome of HF patients with novel-dose S/V therapy, that was, Model 1(age, VT, VHD, COPD, DM and dose) and Model 2 (age, VT, DM and dose).

**Table 1.** Univariate and multivariate competing risk analysis of patients’ characteristics influencing primary outcome (all-cause readmission) based on training dataset

| Characteristics |  | Univariable competing risk analysis | Multivariable competing risk analysis |  |  |
| --- | --- | --- | --- | --- | --- |
|  |  | HR (95%CI, *P*-value) | HR (95%CI, *P*-value)<sup>a</sup> | HR (95%CI, *P*-value)<sup>b</sup> | HR (95%CI, *P*-value)<sup>c</sup> |
| Gender (Male) |  | 0.97 (0.48-1.94, p=0.930) | - | - | - |
| Age | <55y | Reference | Reference | Reference | Reference |
|  | 55-<65y | 0.42 (0.14-1.22, p=0.110) | 0.32 (0.11-0.96, p=0.041) | 0.34 (0.12-0.95, p=0.040) | 0.38 (0.13-1.12, p=0.080) |
|  | 65-<75y | 1.61 (0.73-3.55, p=0.240) | 1.13 (0.50-2.55, p=0.770) | 1.23 (0.54-2.83, p=0.620) | 1.53 (0.71-3.29, p=0.270) |
|  | 75+y | 3.09 (1.21-7.84, p=0.018) | 1.72 (0.55-5.42, p=0.350) | 2.42 (0.94-6.19, p=0.065) | 2.95 (1.18-7.35, p=0.021) |
| BMI | <18.5kg/m<sup>2</sup> | Reference | Reference | - | - |
|  | 18.5-<24kg/m<sup>2</sup> | 0.66 (0.18-2.39, p=0.530) | 1.31 (0.30-5.76, p=0.720) | - | - |
|  | 24-<28kg/m<sup>2</sup> | 0.23 (0.06-0.94, p=0.041) | 0.48 (0.09-2.62, p=0.390) | - | - |
|  | 28+kg/m<sup>2</sup> | 0.41 (0.10-1.60, p=0.200) | 1.26 (0.26-6.16, p=0.770) | - | - |
| DCM (Presence) |  | 0.96 (0.41-2.25, p=0.920) | - | - | - |
| Hypertension (Presence) |  | 1.13 (0.56-2.27, p=0.730) | - | - | - |
| CHD (Presence) |  | 1.13 (0.45-2.83, p=0.800) | - | - | - |
| MI (Presence) |  | 1.51 (0.37-6.27, p=0.570) | - | - | - |
| VHD (Presence) |  | 2.80 (1.25-6.28, p=0.013) | 2.35 (0.94-5.86, p=0.068) | 2.14 (0.89-5.17, p=0.090) | - |
| Bradycardia (Presence) |  | 0.94 (0.28-3.17, p=0.920) | - | - | - |
| PACs (Presence) |  | 2.12 (0.97-4.62, p=0.060) | - | - | - |
| PVCs (Presence) |  | 2.95 (1.57-5.55, p=0.001) | 1.77 (0.75-4.17, p=0.190) | - | - |
| VT (Presence) |  | 4.38 (1.81-10.55, p=0.001) | 2.93 (1.05-8.23, p=0.041) | 3.59 (1.33-9.69, p=0.012) | 3.50 (1.32-9.32, p=0.012) |
| COPD (Presence) |  | 3.45 (1.27-9.38, p=0.015) | 2.92 (0.91-9.42, p=0.073) | 2.70 (0.97-7.51, p=0.056) | - |
| DM (Presence) |  | 2.28 (1.02-5.10, p=0.045) | 3.22 (1.35-7.66, p=0.008) | 2.70 (1.17-6.22, p=0.020) | 2.26 (1.05-4.86, p=0.038) |
| SRI (Presence) |  | 1.93 (0.31-11.86, p=0.480) | - | - | - |
| Dose | ≤50mg | Reference | Reference | Reference | Reference |
|  | 50-<100mg | 0.38 (0.20-0.74, p=0.004) | 0.51 (0.24-1.07, p=0.077) | 0.45 (0.23-0.89, p=0.021) | 0.39 (0.20-0.80, p=0.009) |
|  | 100+mg | 0.14 (0.05-0.38, p<0.001) | 0.24 (0.08-0.77, p=0.016) | 0.21 (0.07-0.59, p=0.003) | 0.16 (0.06-0.45, p<0.001) |

Note: <sup>a</sup> Variables from least absolute shrinkage and selection operator approach and Akaike's Information Criterion, <sup>b</sup> Model 1, <sup>c</sup> Model 2. Abbreviations: BMI, body mass index; DCM, dilated cardiomyopathy; CHD, coronary heart disease; MI, myocardial infarction; VHD, valvular heart disease; PACs, premature atrial contractions; PVCs, premature ventricular contractions; VT, ventricular tachycardia; COPD, chronic obstructive pulmonary disease; DM, diabetes mellitus; SRI, severe renal impairment; HR, hazard ratio; CI, confidence interval.

C-index was 0.752 (95% confidence interval (CI) : 0.724, 0.789) and 0.702 (95%CI: 0.663, 0.743), time-dependent AUC (Figure S3) was more than 0.677 and 0.556 over the follow-up period for Model 1 and 2, respectively. The calibration curves depicted that observed and predicted values of two models were almost consistent (Figure S4). Decision curves were illustrated in Figure S5, and exhibited that clinical net benefit gained from Model 1 was higher than two hypothetical scenarios (none readmission, all readmission) and Model 2 when the threshold probabilities were within the range of 0%-31% and 0%-7% for 30-day and 6-month readmission, respectively. Model 1 also had a wider range of threshold probabilities (0%-10%, 17%-20%, 23%-33%, 39%-50%) in predicting 3-month readmission. Therefore, Model 1 had considerable discriminative and calibrating abilities, and clinical utility, which was displayed as a static nomogram (Figure3) and a web-based dynamic nomogram (https://haoxx.shinyapps.io/DynNom_HF/).

The Nomogram implied that patients aged 75 years and above were at the highest risk of having novel-dose related readmission, followed by those aged <55y, 65-<75y, 55-<65y. Then all-cause readmission mainly occurred in patients with the lowest-dose S/V, followed by the lower- and low-dose. Lastly, other factors associated with an increase in all-cause readmission included history of VT, VHD, COPD and DM.

### Causal mediation effects

An omnibus mediation effect analysis (Figure 4) indicated that LVRR had a significant negative indirect effect on the difference of novel-dose S/V in all-cause readmission, accounting for 20.3% of the total effect. Then the relative mediation effect analysis showed that the odds of having readmission for the lower- and low-dose were respectively on average 0.45 (95% CI: 0.37, 0.56) and 0.14 (95%CI: 0.10, 0.19) times that for the lowest-dose in total effect, while the corresponding odds of indirect effect of LVRR were 0.92 (95%CI: 0.91, 0.94) and 0.978 (95%CI: 0.971, 0.984) times.

**Figure 4.**
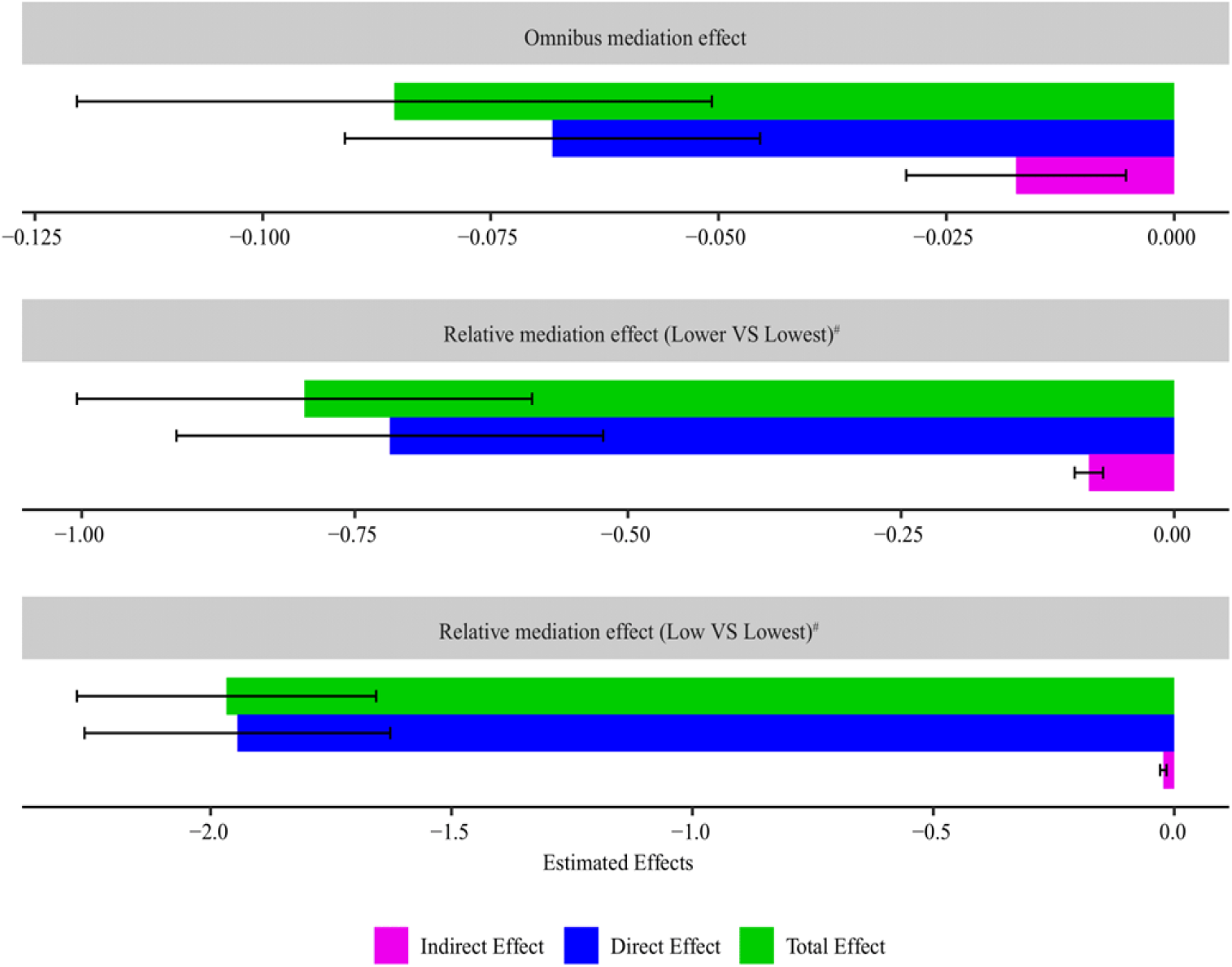
Estimated mediation effects (with 95% confidence intervals) of left ventricular reverse remodeling on the association between novel-dose Sacubitril/Valsartan (S/V) and hospital readmission of patients with heart failure. Note: Predictor variable (S/V dosage) was continuous and categorical in omnibus and relative mediation effect analysis. Black line with error bars represented the 95% confidence intervals corresponding to quantile estimators from the bootstrap. Additionally, total effect = indirect effect + direct effect (see Figure S1). # Lowest-dose S/V was the reference.

## Discussion

Although S/V has been recommended in clinical practice guidelines for patients with HFrEF, some were only able to tolerate novel dosage (below the standard) dominantly due to hypotension. Our study was an attempt to assess the effects of novel-dose S/V on hospital readmissions in these patients, establish clinical prediction models, as well as explore the possible mediation effect of LVRR. Berg et al stated that the efficacy and safety of different S/V levels seemed to be consistent no matter whether patients obtained the maximum dose^34^. Consistently, our study showed significantly different cumulative incidences of all-cause readmission. The risk of all-cause readmission for patients with lowest-dose S/V were much higher than those with the lower- and low-dose, and mostly occurred within 1 year. However, in our small sized cohort, no difference in all-cause death among the three novel-dose S/V were observed. The clinical effects of novel-dose S/V remained to be further investigated.

Reducing hospital readmission was one of the primary targets of the HF treatment. However, it remained unsolved that how to predict which patients with HF would suffer hospital readmission. In this ARNI therapy cohort, we built a nomogram model, which revealed that age, VT, VHD, COPD, DM and dose were associated with an increased risk of all-cause readmission. As expected, patients who taken the lowest-dose S/V were at the highest risk of all-cause readmission, followed by those who taken lower- and low-dose, which was in accordance with cumulative incidence curves. Previous studies have reported some factors associated with hospital readmission of HF patients, such as, male gender^35^, age^36^, DM^36^, VT^37^, VHD^38^, COPD^35,39^, renal dysfunction^40^, psychiatric illness^40^, etc. Similarly, our study suggested that patients aged 75+y were the high-risk population, followed by those aged <55y, 65-<75y, 55-<65y. The elderly patients were mostly observed to have low degrees of physical activity and depression and anxiety, as well as concomitant chronic diseases, which were likely to result in increasing disease risk and aggravating clinical outcomes^41,42^. Whellan et al. pointed out a dichotomous relationship between age and risk of death or readmission, namely, risk of readmission or death decreased as age increased up to 55 years of age, then increased in those older than 55 years^43^.

Thus, there might be a tick-shaped relationship between age and all-cause readmission in patients taking novel-dose. Future studies might help more finely identify the high-risk population. In addition, patients with VT, or COPD, or DM were at significantly higher risk of all-cause readmission. This might indicate that targeted treatments of these comorbidities would provide a potential opportunity to improve outcomes in patients with HF.

Reverse cardiac remodeling refers to the recovery of ventricular function and reduction in cardiac volumes. Reverse remodeling has become a primary objective in HF treatment. Guideline directed medical and device therapies has been proven to result in the reverse cardiac remodeling. Numerous studies have highlighted the role for RAAS blockade in promoting reverse remodeling in patients with HFrEF. The SOLVD and Val-HeFT studies have demonstrated the beneficial effects of ACEI and ARB on LVRR^44-47^, respectively. Since the landmark of PARADIGM-HF study^4^, the data support the link between ARNI and LVRR was growing. A meta-analysis including more than 10,000 population showed that ARNI has significant improvement over ACEI/ARB on the left LV dimensions and EF^48^. The EVALUATE-HF and PROVE-HF trial both observed the reduction of multiple atrial and ventricular parameters of remodeling after initiation of ARNI ^49,50^. However, the relations between ARNI dosage and the LVRR are not addressed. In our study, we classified the patients into different dosage group and revealed that the under target novel dose remained effective on LVRR.

Mechanistically, LVRR might be a part of the explanation of reduced readmission. The mediation effects of a variety of covariates on all-cause readmission was explored. The omnibus mediation effect analysis revealed that novel-dose S/V was negatively related to all-cause readmission through its effect on LVRR, which has been identified as a critical mediator accounting for approximately 20.3% of total effect. Meanwhile, our study demonstrated that the lower- and low-dose S/V decreased the risk of readmission by 55% (1-0.45) and 86% (1-0.14) as compared with the lowest-dose, respectively, whereas only on average 8% (1-0.92) and 2.2% (1-0.978) of the readmission risk were correspondingly reduced through the mediation of LVRR. Although LVRR had a significant negative indirect effect on the difference of novel-dose S/V in all-cause readmission, the relative effect caused by LVRR was similar between the lower- (or low-) and lowest-dose levels. It seemed that the novel-dose S/V might exert its effect through other undermined mechanisms.

There were several limitations with our study. First, as a single-center retrospective study, our study performed internal validation for the prediction models, which still needed further external validation using an independent cohort. Second, VHD and COPD were marginally significant variables in our proposed Model 1, probably because the sample size was relatively small. Future studies with a larger sample size would be required to confirm the model.

## Conclusion

Our study developed a clinically useful competing-risk nomogram for predicting all-cause readmission in HF patients receiving novel-dose S/V therapy. The findings revealed that the novel-dose S/V, especially the lowest, significantly affected the hospital readmissions, in part mediated by LVRR. However, the clinical benefits of different novel-dose S/V and more potential novel mechanisms still required further in-depth examination.

## Data Availability

Data not available due to ethical restrictions.

https://haoxx.shinyapps.io/DynNom_HF/

## Supplementary Material

The Supplementary Material for this article can be found online as additional file 1.

## Acknowledgments

We would like to thank Lili Yang for her technical assistance in the data search and process.

## Authors’ contributions

Zhexue Qin Xiaolin Luo and Xi Liu were involved in conception and design. Changchun Hou, Ling Chen, and Juhao Yang carried out the acquisition of data. Ning Sun, Enpu Yang, Zebi Wang, Yun Cui, Jing Zhong and Luyu Wang contributed to the follow up of the patients. Xinxin Hao and Changchun Hou performed the statistical analysis and drafted the manuscript. Xiaolin Luo and Zhichun Gao contributed to interpretation of data. All authors contributed to critical revision of the manuscript for important intellectual content. Xinxin Hao, Changchun Hou, Xi Liu and Zhexue Qin were involved in final revision of manuscript. All authors read and approved the final manuscript.

## What is new?

The study was the first attempt to explore the effects of the novel-dose Sacubitril/Valsartan (S/V) on all-cause readmission in patients with heart failure (HF) using the competing-risk analysis, as well as investigate the possible mediator role of left ventricular reverse remodeling (LVRR).

A nomogram based on the competing-risk regression model was established to predict all-cause readmission of patients who taken novel-dose S/V. Moreover, our study was useful in gaining a better understanding of the relationship and mediating mechanism between novel-dose S/V, LVRR, all-cause readmission.

## What are the clinical implications?

So far there was little data on the novel-dose S/V in a real world setting. The findings of our study would provide new insights on the effects of novel-dose S/V, as well as useful information for health decision-making.

